# TGF-β1 in extracellular vesicles from HIV-infected plasma and macrophages linked to cardiopulmonary dysfunction

**DOI:** 10.1101/2020.09.09.20191338

**Authors:** Balaji Krishnamachary, Stuti Agarwal, Aatish Mahajan, Ashok Kumar, Aradhana Mohan, Ling Chen, Priscilla Hsue, Prabhakar Chalise, Alison Morris, Navneet K. Dhillon

## Abstract

**Rationale:** Extracellular vesicles (EVs) have emerged as important mediators in cell-cell communication and disease pathogenesis; however, their relevance in pulmonary hypertension (PH) secondary to HIV infection is yet to be explored.

**Objective:** To examine the role of circulating small EVs and monocyte-derived macrophage (MDM) EVs in the development of HIV-associated PH

**Methods:** EVs isolated from plasma of HIV-infected drug users and non-users with/without PH and from supernatants of HIV-infected MDMs treated with/without second hit of cocaine were studied for their effect on vascular dysfunction both *in vitro* and *in vivo*.

**Measurements and Main Results:** We report significantly higher numbers of plasma derived EVs (PEVs) carrying higher levels of TGF-β1 in people living with HIV (PLWH) that had PH compared to non-PH PLWH. Importantly, levels of these TGF-β1 loaded PEVs correlated with pulmonary arterial systolic pressures, CD4 counts, but not with diffusion capacity for carbon monoxide or viral load. Correspondingly, enhanced TGF-β1-dependent pulmonary endothelial injury and smooth muscle hyperplasia was observed. Cocaine treatment of HIV-1 infected-MDMs resulted in increased number of TGF-β1 high-EVs. Intravenous injection of these EVs in rats led to increased right ventricle systolic pressure accompanied with myocardial injury and increased levels of serum endothelin-1, TNF-α, and cardiac Troponin-I. Conversely, pretreatment of rats with TGFβ-Receptor 1 inhibitor prevented these EV-mediated changes.

**Conclusion:** Findings define the ability of macrophage-derived small EVs to cause pulmonary vascular modeling and PH via modulation of TGF-β signaling and suggest clinical implications of circulating TGF-β high-EVs as a potential biomarker of HIV-PH.

## INTRODUCTION

Pulmonary arterial hypertension (PAH) is one of the most devastating non-infectious complications related to HIV infection and echocardiographic evidences suggest prevalence of increased pulmonary artery systolic pressure (PASP) in 2.6–15.5% of people living with HIV (PLWH) (1), (2). HIV-associated PAH falls under Group I pulmonary hypertension (PH) and the risk factors such as cocaine, methamphetamine and intravenous drug use (IVDU) in PLWH can potentiate the development of HIV-PAH (2-4). Furthermore, with the increasing burden of cardiac and chronic respiratory complications in PLWH, the prevalence of secondary PH is suggested to increase as well, in these individuals (5, 6).

The presence of interstitial and perivascular infiltration of inflammatory cells has been linked to intimal and medial thickening of blood vessels associated with PH (7, 8). Recently, we reported increased perivascular macrophages in the remodeled thickened vessels in the lung sections from HIV-infected humans, simian immunodeficiency virus (SIV)-infected macaques and HIV-transgenic (HIV-Tg) rats, particularly in the presence of illicit drug exposure (9, 10). However, the extent to which the perivascular inflammation is involved in the thickening and occlusion of blood vessels is elusive. It is believed that inflammation precedes vascular remodeling in PAH and is more often an origin rather than a result of PAH (7, 8). Circulating monocytes contribute to increased interstitial macrophages observed around the remodeling pulmonary vessels in PH (11, 12).

Other than the direct release of viral proteins, cytokines and growth factors to the vascular cells, immune cells may also deliver their molecular cargo through the release of extracellular vesicles (13, 14). Extracellular vesicles (EVs) are small membrane vesicles ranging from 30–1000 nm and are involved in intercellular communication by delivering protein/nucleic acid cargo to surrounding cells. Studies have emphasized upon changes in the EV cargo to the development of various diseases in animal models including PAH (15, 16). However, limited studies have associated the levels of circulating membrane vesicles with pulmonary vascular resistance in PAH patients (17, 18). These studies were focused on larger micro-particles and lacked the details on type, size and composition of EVs, leaving open the question of the role of small EVs.

Recent reports suggest the presence of viral proteins in the bronchoalveolar lavage fluid -
and plasma -derived EVs from PLWH (19, 20) (21). Given the importance of TGF-β signaling in the development of PH (22, 23) (24) and recent reports showing the presence of TGF-β1 on the surface and within exosomes (25), we hypothesize that the increased levels of TGFβ-loaded EVs in the plasma from PLWH, may associate with the presence of PH and that EVs released by HIV-infected monocyte-derived macrophages to cause the pulmonary vascular changes and right ventricle dysfunction in wild-type and non-infectious HIV-Tg rats.

## MATERIAL AND METHODS

Detailed description of methods and statistical analyses is provided in the online supplement.

## RESULTS

### Plasma-derived EVs (PEVs) are elevated in HIV infected and /or cocaine users

Number of EVs were compared in the plasma samples from HIV-infected individuals with (PLWH+Coc, n = 15) or without history of cocaine abuse (PLWH, n = 15) and uninfected individuals with the history of cocaine abuse (Coc, n = 15) and non-drug users (UI, n = 15) (Table 1). Gender, race and CD4 cell count were the only variables observed to be significantly different among groups(Table 1). NanoSight analysis showed highest total number of EVs in the plasma from PLWH-cocaine users when compared with all other groups (Fig. 1A-B) with significant higher numbers of EVs observed in the size-range of 100–250 nm (Fig. 1C). These small EVs were further characterized by TEM demonstrating classic lipid bi-layered structures (Fig. 1D) and by western blot showing the presence of exosomal markers (Fig. 1E).

**Table 1:**
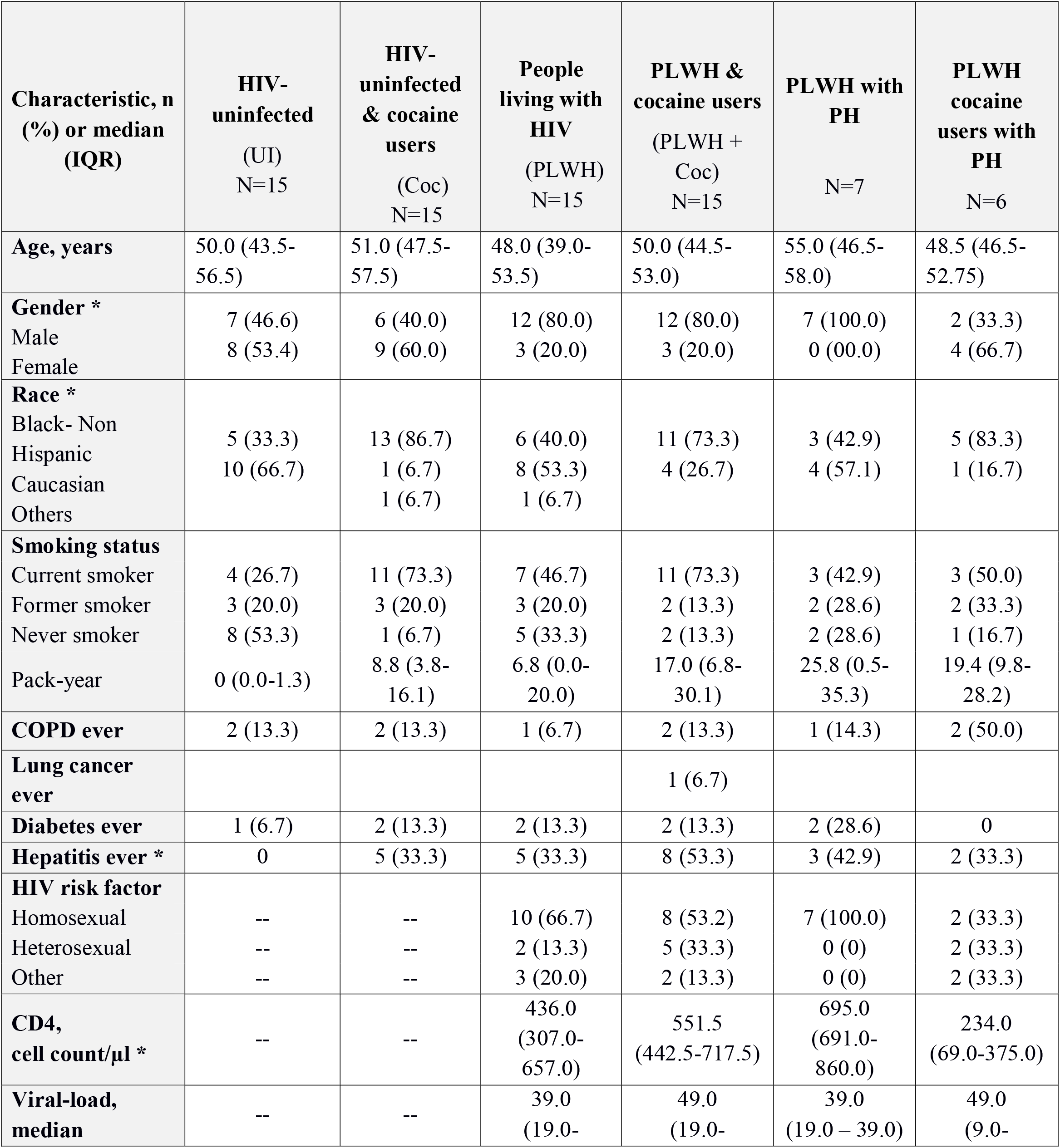
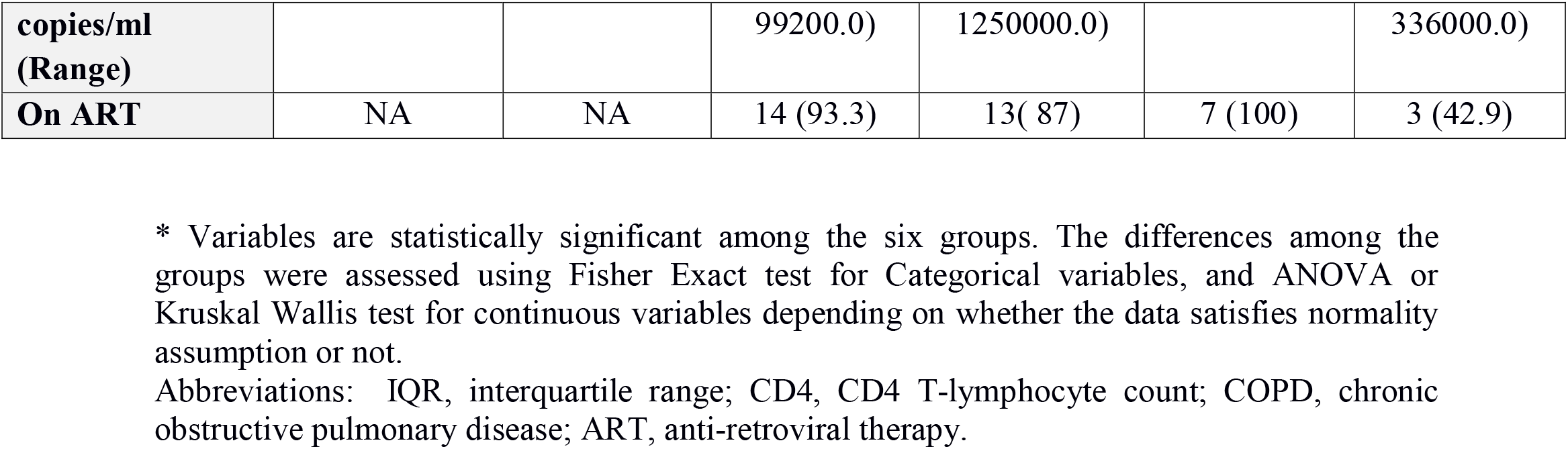
Demographic and clinical characteristics of participants by HIV status and cocaine use.

**Figure 1:**
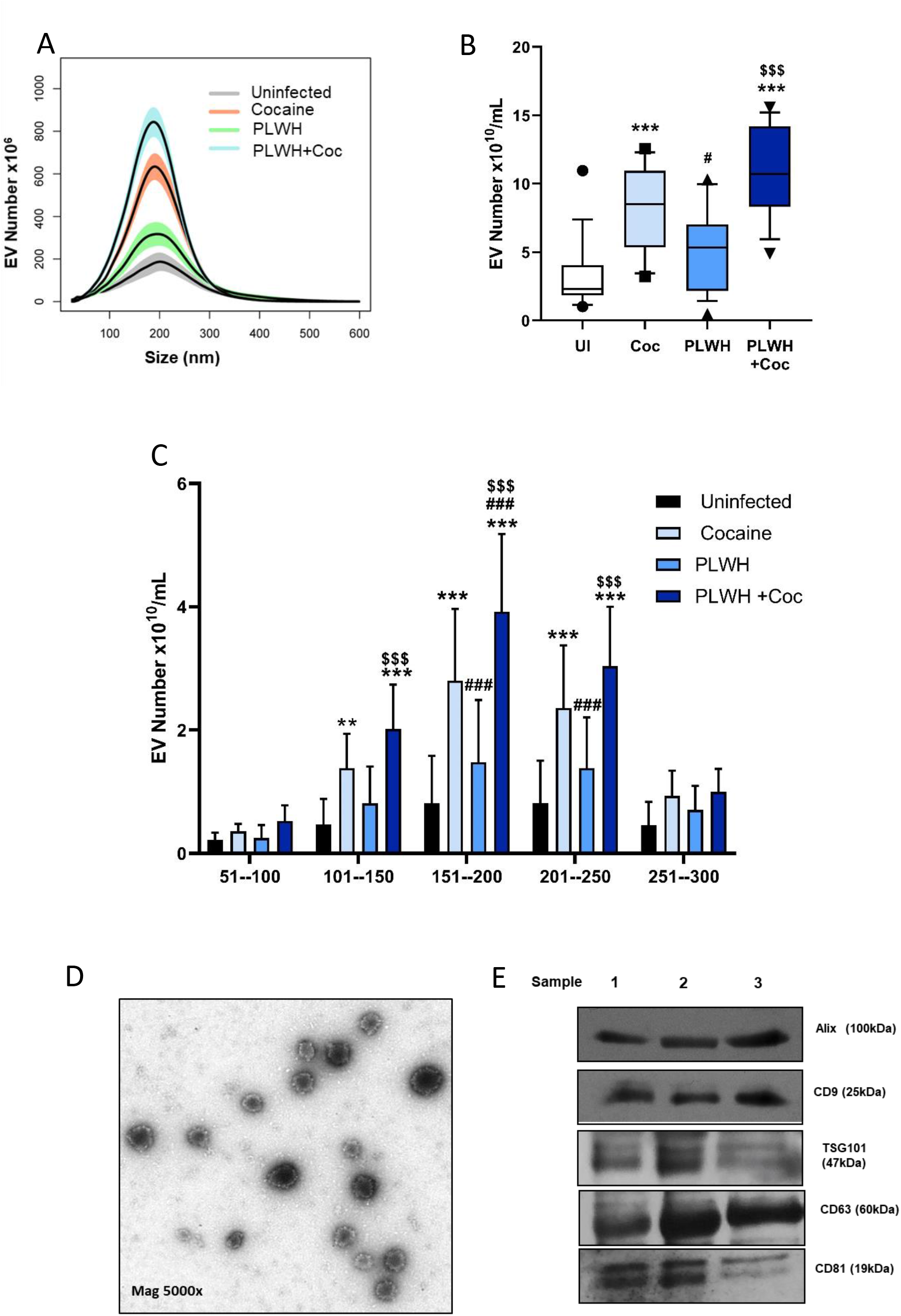
Analysis of plasma derived extracellular vesicles (PEVs) from HIV-infected and/or cocaine treated individuals. Extracellular vesicles were isolated from 0.5ml of EDTA-plasma of HIV-uninfected healthy controls (UI), HIV-uninfected cocaine users (Coc), people living with HIV (PLWH) non drug users and PLWH cocaine users (PLWH+Coc) (n = 15/group) using exoEasy kit. **A-C**, Particles were counted and characterized for size distribution by NanoSight nanoparticle tracking analysis. Data represents number of EVs/ml of final EV suspension obtained from 0.5ml EDTA plasma using exoEasy kit. Boxes in B panel depicts median and IQR, whiskers show 10–90 percentiles. **D-E**, Characterization of PEVs by Transmission electron microscopy (TEM) and western blot analysis. Representative TEM image of PEVs at 5000X magnification (C) and western blot of PEV protein extract from three different subjects for exosomal markers (D). ***p< 0.001, *p< 0.05 vs. UI, # p< 0.05, ##p< 0.01, ###p< 0.001 vs. Coc, $$$ p< 0.001 vs. PLWH.

### PEVs from HIV-infected and/or cocaine users promote TGF-β dependent pulmonary endothelial and smooth muscle dysfunction

EVs are readily taken up by human pulmonary arterial smooth muscle cells (HPASMCs) (21) as well as by human pulmonary microvascular endothelial cells (HPMECs) (Supplementary Figure IIA). Addition of PLWH+Coc or Coc group PEVs on HPMECs for 24h resulted in significant increased apoptosis when compared to the treatment with an equal amount of EVs from UI controls (Fig. 2A). Alternatively, significant increased proliferation of HPMECs and HPASMCs was observed in response to 48h treatment with PEVs from PLWH+Coc, PLWH or Coc when compared with the cells loaded with UI PEVs (Fig. 2B & C). This increased proliferation of both cell-types on treatment with PLWH+Coc EVs was higher when compared to PLWH EVs, although not significant. Nevertheless, a significantly higher number of EVs observed in the plasma of PLWH+Coc group (10.9 × 10^6^ ± 3.2/0.5ml) compared to PLWH-group plasma (5.3 × 10^6^ ± 2.8/0.5ml) (Fig. 1B) is expected to have more cumulative effect on the cellular injury *in-vivo*.

**Figure 2:**
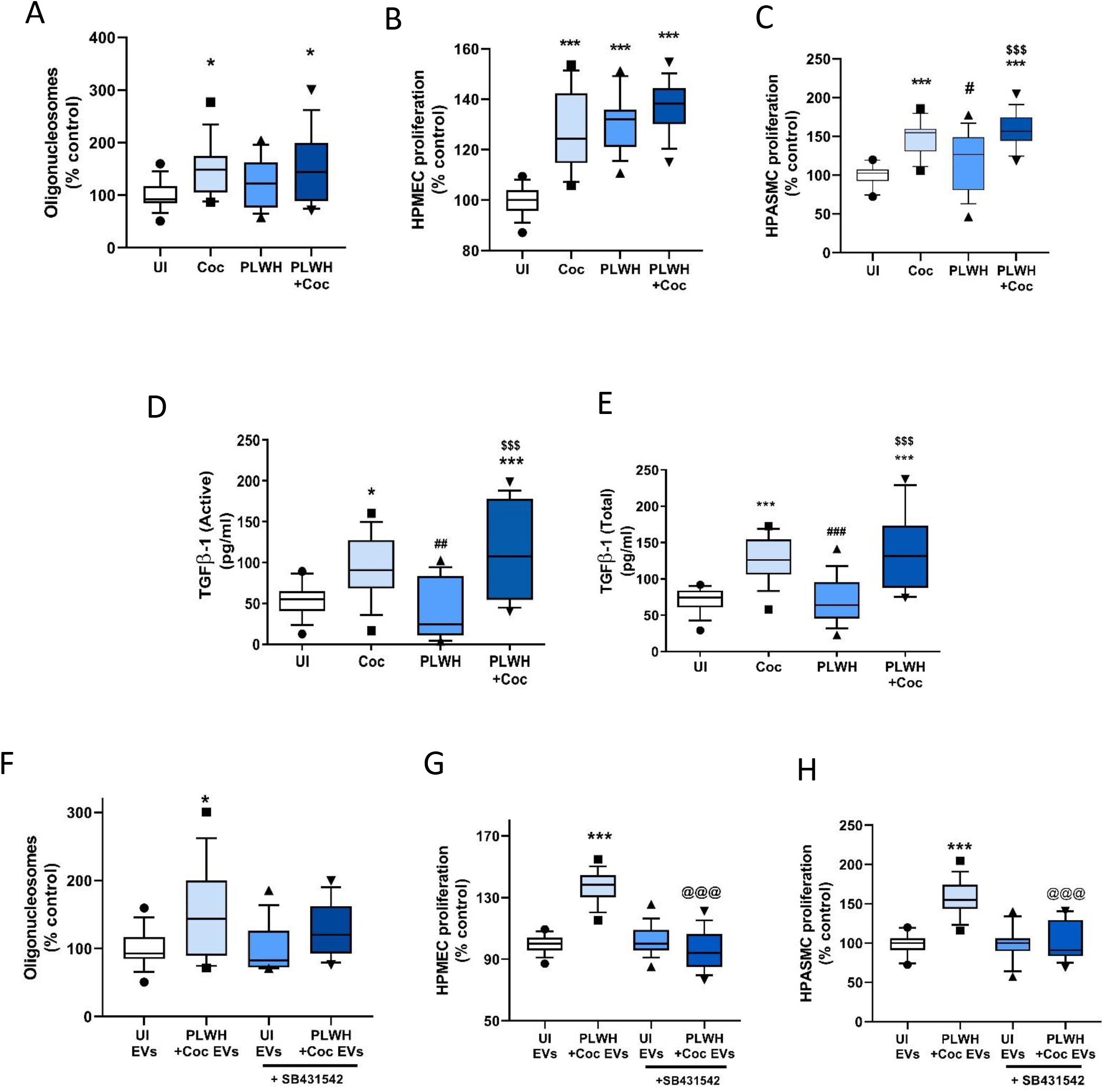
Increased levels of TGF-β1 in plasma-derived EVs from PLWH cocaine users promote pulmonary endothelial and smooth muscle dysfunction. Analysis of endothelial apoptosis (**A**) and proliferation (**B)** on treatment with PEVs from HIV- uninfected (UI), HIV-uninfected cocaine users (Coc), people living with HIV (PLWH) non drug users and PLWH cocaine users (PLWH+Coc) (n = 15/group). Human pulmonary microvascular endothelial cell (HPMEC) were plated in 96-well plate, serum starved after 24h in 0.5% serum containing media followed by addition of 3 µg of PEVs/well and analysis of apoptosis using Cell death ELISA after 24h (A) and cell proliferation assay after 48h (B). **C**, Analysis of human pulmonary arterial smooth muscle cell (HPASMC) proliferation on treatment with PEVs. After 72h of plating in 96-well plate, cells were serum starved for 48h, followed by addition of 3 µg of PEVs/well. MTS cell proliferation assay was performed at 48h post-treatment. **D-E**, Plasma derived EVs (PEVs) were analyzed for active (D) and total (E) TGFβ-1 levels by ELISA. **F**, Cell death ELISA on HPMECs treated with PLWH+Coc PEVs in the presence or absence of 10µM of SB431542 (TGFβ-R1 inhibitor). **G-H**, MTS proliferation assay was performed on HPMECs (G) and HPASMCs (H) after 48h treatment with PLWH+Coc PEVs in the presence or absence of SB431542. n = 15/group were used for all experiments Boxes represent median and IQR, whiskers show 10–90 percentiles. ***p< 0.001,*p< 0.05, vs UI; #p< 0.05,##p< 0.01, ### p< 0.001 vs Coc; $$$p< 0.001 vs PLWH and @@@ p< 0.001 vs PLWH+Coc.

Analysis of TGF-β1 levels in PEVs showed significant increased levels of active (Fig. 2D) as well as total (active plus latent) (Fig. 2E). TGF-β1 in PEVs from HIV-uninfected cocaine user (Coc) and PLWH+Coc groups when compared to PEVs from UI controls. Interestingly, levels of TGF-β1 in PLWH-PEVs were same as observed in UI-PEVs (Fig. 2D-E) and higher levels in PEVs from PLWH+Coc group were also significant compared to PLWH group. Treatment of cells in the presence of TGFβ-Receptor 1 inhibitor: SB431542 (10µM) could significantly rescue the PLWH+Coc PEV-mediated enhanced endothelial as well as smooth muscle cell proliferation (Fig. 2G and H) but didn’t show any significant effect on PEV-mediated EC apoptosis (Fig. 2F).

### Significant increased TGF-β1 levels in PEVs from PLWH that had PH compared to non-PH individuals

To assess the alterations in EV numbers and associated TGF-β1 levels in PLWH with PH, we compared PLWH (n = 13) cocaine users or non-users that had PH with PLWH (n = 30) and HIV-un-infected (n = 30) subjects with and without history of cocaine use that had no PH (Table 1 & 2). PEVs from PLWH+PH subjects were significantly higher in number compared to PLWH without PH (Fig. 3A), even after adjusting for gender and race (Table 2). Although, PLWH-PH PEVs showed no significant difference on apoptosis of HPMECs when compared to EVs from PLWH without PH (Fig. 3Bi), augmented proliferation of endothelial and smooth muscle cells was observed with PLWH-PH PEVs (Fig. 3Bii and C). Interestingly, presence of PH among PLWH was associated with a significantly higher levels of both active and total TGFβ-1 in PEVs (Fig. 3D) before and after adjusting for covariates (Table 2). Increased EC and SMC proliferation by EVs from PLWH with PH was also found to be TGFβ dependent (Supplementary figure I A-C).

**Table 2:**
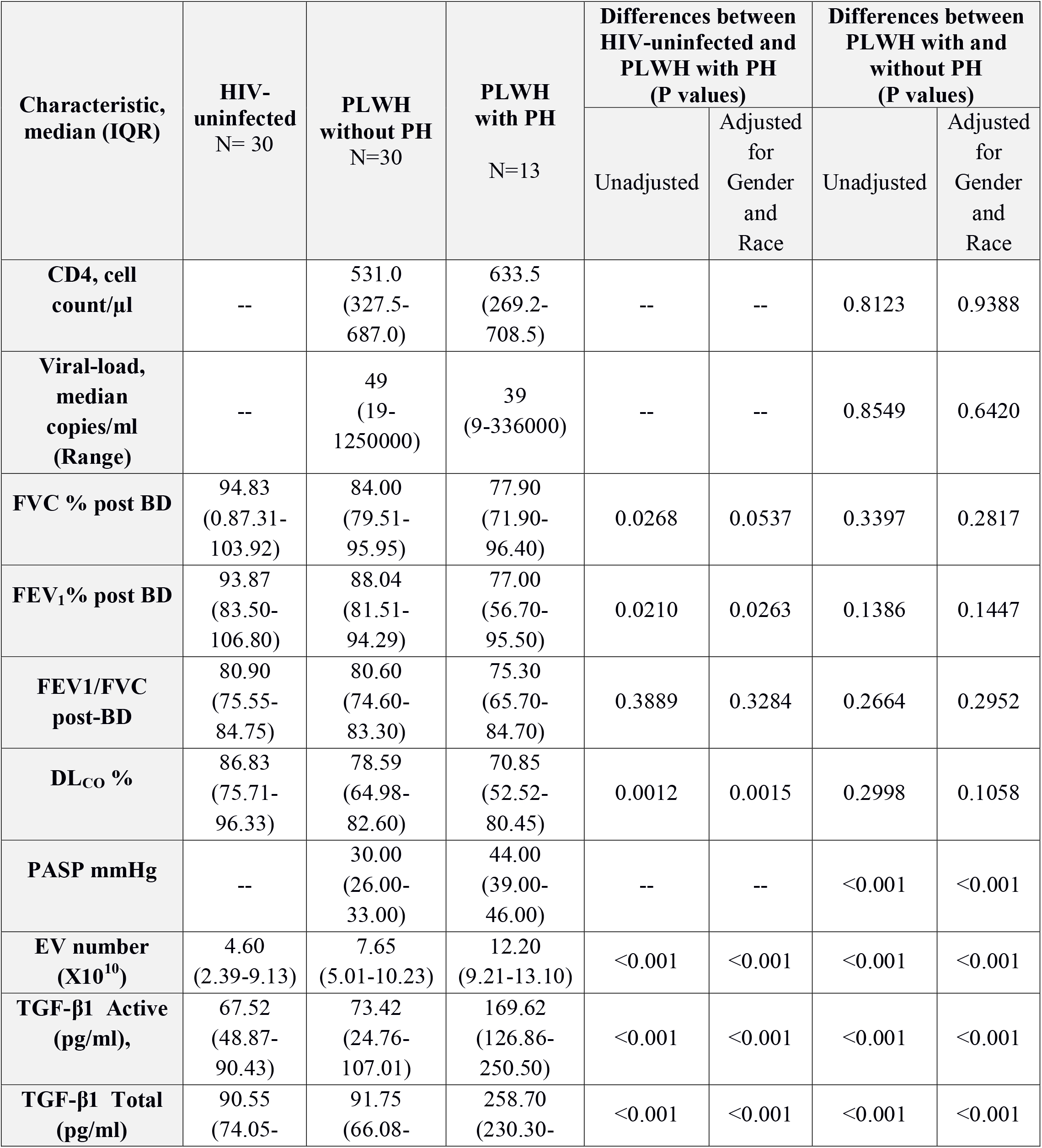
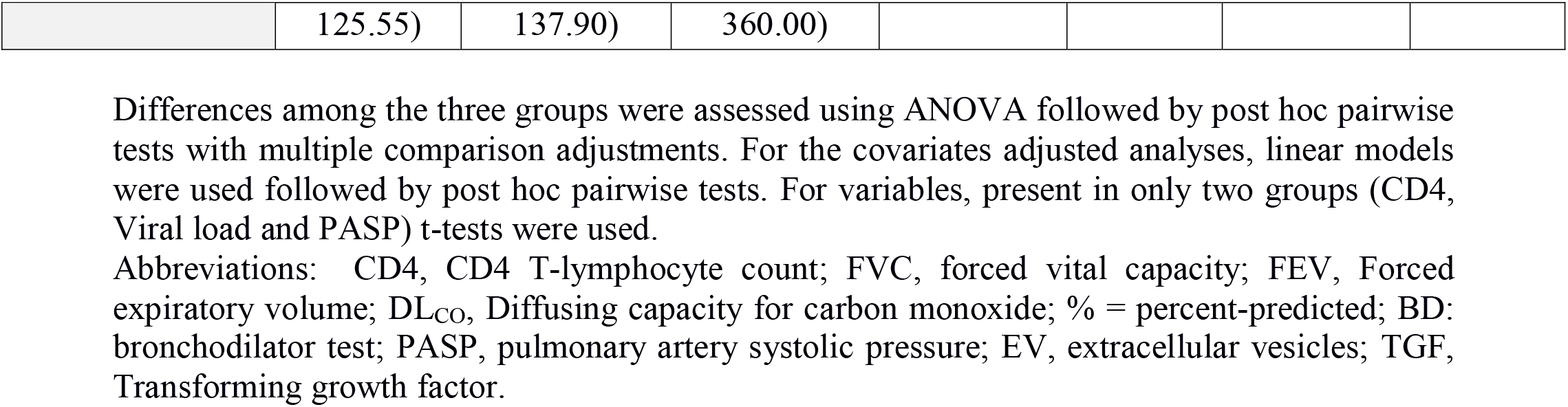
Comparison of lung function, pulmonary arterial systolic pressures (PASP), EV number and TGF-β (active and total) levels in HIV-uninfected or PLWH- without PH and PLWH with PH.

| Characteristic,<br>median (IQR) | HIV-uninfected<br>N= 30 | PLWH<br>without PH<br>N=30 | PLWH<br>with PH<br>N=13 | Differences between<br>HIV-uninfected and<br>PLWH with PH<br>(P values) |  | Differences between<br>PLWH with and<br>without PH<br>(P values) |  |
| --- | --- | --- | --- | --- | --- | --- | --- |
|  |  |  |  | Unadjusted | Adjusted<br>for<br>Gender<br>and<br>Race | Unadjusted | Adjusted<br>for<br>Gender<br>and<br>Race |
| **CD4, cell**<br>**count/μl** | -- | 531.0<br>(327.5-687.0) | 633.5<br>(269.2-708.5) | -- | -- | 0.8123 | 0.9388 |
| **Viral-load,**<br>**median**<br>**copies/ml**<br>**(Range)** | -- | 49<br>(19-1250000) | 39<br>(9-336000) | -- | -- | 0.8549 | 0.6420 |
| **FVC % post BD** | 94.83<br>(0.87.31-103.92) | 84.00<br>(79.51-95.95) | 77.90<br>(71.90-96.40) | 0.0268 | 0.0537 | 0.3397 | 0.2817 |
| **FEV<sub>1</sub>% post BD** | 93.87<br>(83.50-106.80) | 88.04<br>(81.51-94.29) | 77.00<br>(56.70-95.50) | 0.0210 | 0.0263 | 0.1386 | 0.1447 |
| **FEV1/FVC**<br>**post-BD** | 80.90<br>(75.55-84.75) | 80.60<br>(74.60-83.30) | 75.30<br>(65.70-84.70) | 0.3889 | 0.3284 | 0.2664 | 0.2952 |
| **DL<sub>co</sub> %** | 86.83<br>(75.71-96.33) | 78.59<br>(64.98-82.60) | 70.85<br>(52.52-80.45) | 0.0012 | 0.0015 | 0.2998 | 0.1058 |
| **PASP mmHg** | -- | 30.00<br>(26.00-33.00) | 44.00<br>(39.00-46.00) | -- | -- | <0.001 | <0.001 |
| **EV number**<br>**($\times 10^{10}$)** | 4.60<br>(2.39-9.13) | 7.65<br>(5.01-10.23) | 12.20<br>(9.21-13.10) | <0.001 | <0.001 | <0.001 | <0.001 |
| **TGF-β1 Active**<br>**(pg/ml),** | 67.52<br>(48.87-90.43) | 73.42<br>(24.76-107.01) | 169.62<br>(126.86-250.50) | <0.001 | <0.001 | <0.001 | <0.001 |
| **TGF-β1 Total**<br>**(pg/ml)** | 90.55<br>(74.05- | 91.75<br>(66.08- | 258.70<br>(230.30- | <0.001 | <0.001 | <0.001 | <0.001 |
|  | 125.55) | 137.90) | 360.00) |  |  |  |  |

Differences among the three groups were assessed using ANOVA followed by post hoc pairwise tests with multiple comparison adjustments. For the covariates adjusted analyses, linear models were used followed by post hoc pairwise tests. For variables, present in only two groups (CD4, Viral load and PASP) t-tests were used.
Abbreviations: CD4, CD4 T-lymphocyte count; FVC, forced vital capacity; FEV, Forced expiratory volume; DL<sub>CO</sub>, Diffusing capacity for carbon monoxide; % = percent-predicted; BD: bronchodilator test; PASP, pulmonary artery systolic pressure; EV, extracellular vesicles; TGF, Transforming growth factor.

**Figure 3:**
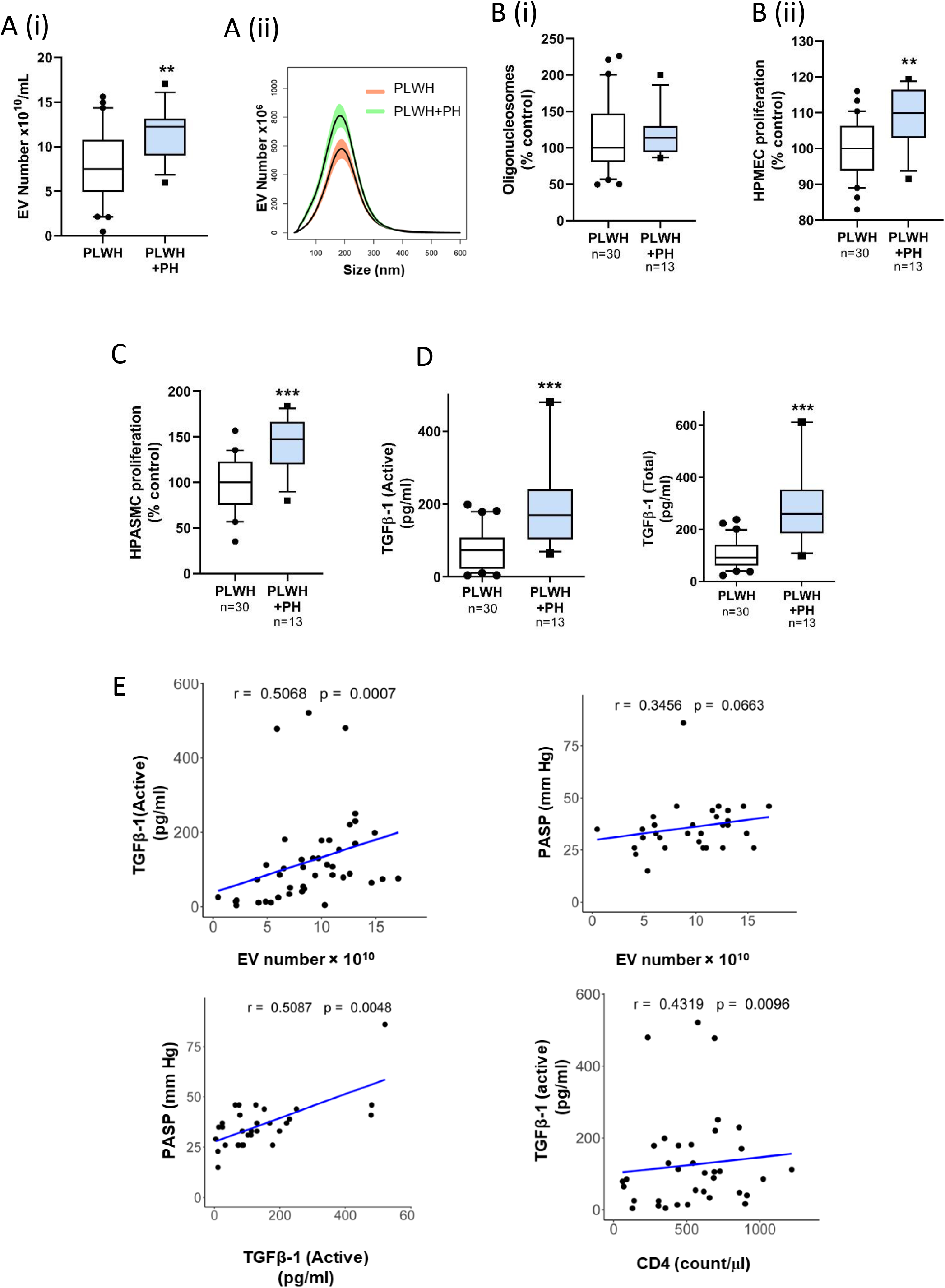
TGF-β1 levels in plasma derived EVs correlate with the presence of PH in HIV- infected individuals. **A**, Size distribution **(i)**, and total number **(ii)** of PEVs were compared between PLWH without PH (n = 30) and with PH (n = 13) by NanoSight. For this, PEVs from PLWH non drug users (n = 15) and PLWH cocaine users (n = 15) without PH were grouped as PLWH group (n = 30). For PLWH PH group (n = 13) plasma samples from PLWH non drug users with PH (n = 8) and PLWH cocaine users with PH (n = 7) were included (Table 1). **B**, To compare the effect of PEVs from PLWH with PH and without PH on the survival and proliferation of endothelial cells, HPMECs were treated with PEVs from both groups (n = 13–15/group) followed by cell death ELISA (i) and MTS assay (ii) at 24h and 48h post-treatment. **C**, Cell proliferation analysis of quiescent HPASMCs treated with PEVs from PLWH with PH and without PH at 48h post-treatment. **D**, Levels of active and total TGFβ-1 in PEVs as analyzed by ELISA. **E**, Correlation analysis of EV numbers with the levels of TGFβ1 (active) and PASP; and of TGFβ1 (active) with PASP and CD4 cell count. For box and whisker plots boxes represent median and IQR, whiskers show 10- 90 percentile. ***p< 0.001, **p< 0.01 vs. PLWH.

While the EV number and TGF-β1 levels in PLWH with PH were also significantly high when compared with UI controls, not much difference was observed between PLWH non-PH and UI groups (Supplementary Table I). Importantly, a significant positive correlation of both active and total isoforms of TGF-β levels in EVs was observed with PASP but not with DL_CO_ in PLWH (Fig. 3E, Supplementary fig. I-D and Table 3). Furthermore, significant association of TGF-β1 level in EVs was observed with CD4 cell count but not with viral load (Fig. 3E, Supplementary fig. I-D and Table 3).

**Table 3:** Correlation analyses of clinical characteristics with EV number and TGF β levels (total and active) in PLWH and HIV-uninfected individuals.

|  | EV number |  | TGF-active |  | TGF total |  |
| --- | --- | --- | --- | --- | --- | --- |
|  | HIV-uninfected<br>N=30 | PLWH<br>N=43 | HIV-uninfected<br>N=30 | PLWH<br>N=43 | HIV-uninfected<br>N=30 | PLWH<br>N=43 |
|  | Correlation<br>(P value) | Correlation<br>(P value) | Correlation<br>(P value) | Correlation<br>(P value) | Correlation<br>(P value) | Correlation<br>(P value) |
| **Age, years** | 0.0154<br>(0.9357) | 0.2284<br>(0.1407) | -0.0036<br>(0.9851) | 0.3061<br>(0.0459) | -0.0188<br>(0.9213) | 0.3241<br>(0.0340) |
| **Caucasian** | -0.5874<br>(0.0006) | -0.0978<br>(0.5328) | -0.5155<br>(0.0036) | -0.1303<br>(0.4049) | -0.5115<br>(0.0039) | -0.1380<br>(0.3776) |
| **Gender** | -0.0350<br>(0.8544) | -0.1043<br>(0.5058) | -0.2448<br>(0.1923) | -0.2041<br>(0.1894) | -0.2565<br>(0.1713) | -0.1508<br>(0.3343) |
| **Pack-year** | 0.2855<br>(0.1489) | 0.3166<br>(0.0386) | 0.1580<br>(0.4313) | 0.3532<br>(0.0201) | 0.2563<br>(0.1970) | 0.3346<br>(0.0283) |
| **Cocaine usage** | 0.7279<br>(5.2e-6) | 0.1495<br>(0.3388) | 0.6123<br>(3.2e-4) | 0.1416<br>(0.3652) | 0.7742<br>(5.2e-7) | 0.0747<br>(0.6340) |
| **CD4 cell count/μl \*** | -- | 0.0868<br>(0.6200) | -- | 0.4319<br>(0.0096) | -- | 0.3802<br>(0.0243) |
| **Viral-load copies/ml \*** | -- | 0.1507<br>(0.3666) | -- | -0.1056<br>(0.5281) | -- | -0.1514<br>(0.3641) |
| **FVC % post BD \*** | 0.2546<br>(0.1911) | 0.2420<br>(0.1325) | 0.1701<br>(0.3869) | 0.1032<br>(0.5264) | 0.0714<br>(0.7182) | 0.0790<br>(0.6280) |
| **FEV1% post BD \*** | 0.3930<br>(0.0386) | 0.0203<br>(0.9009) | 0.1276<br>(0.0.5177) | 0.0110<br>(0.9462) | 0.0959<br>(0.6271) | -0.0010<br>(0.9950) |
| **FEV1/FVC post-BD \*** | 0.3039<br>(0.1159) | -0.4951<br>(0.0012) | -0.1771<br>(0.3674) | -0.3723<br>(0.0180) | -0.0767<br>(0.6981) | -0.3421<br>(0.0307) |
| **DL<sub>CO</sub> % \*** | -0.0099<br>(0.9608) | -0.3515<br>(0.0305) | 0.3636<br>(0.0623) | -0.2978<br>(0.0694) | 0.2883<br>(0.1447) | -0.2881<br>(0.0794) |
| **PASP, mmHg \*** | -- | 0.3456<br>(0.0663) | -- | 0.5087<br>(0.0048) | -- | 0.6189<br>(0.0003) |
| **EV number (X10<sup>10</sup>) \*** | -- | -- | 0.2948<br>(0.1277) | 0.5068<br>(0.0007) | 0.3637<br>(0.0571) | 0.5307<br>(0.0004) |

\*Spearman's rank correlation analysis was adjusted for gender and race.

### Increased TGF-β1 levels in EVs released by HIV-infected monocyte-derived macrophages on exposure to cocaine correspond to augmented endothelial and smooth muscle dysfunction

In light of recent findings demonstrating circulating blood monocytes as a source of perivascular macrophages surrounding the remodeled vessels in PH (7, 8, 11) we next examined the levels of TGF-β1 in EVs released by HIV-infected monocyte derived macrophages (MDM) in the presence and absence of cocaine. Similar to plasma EVs, we observed significant increase in both active and total forms of TGFβ-1 in the EVs from supernatants of HIV-infected MDMs on cocaine treatment (H+C) (Fig. 4A).

**Figure 4:**
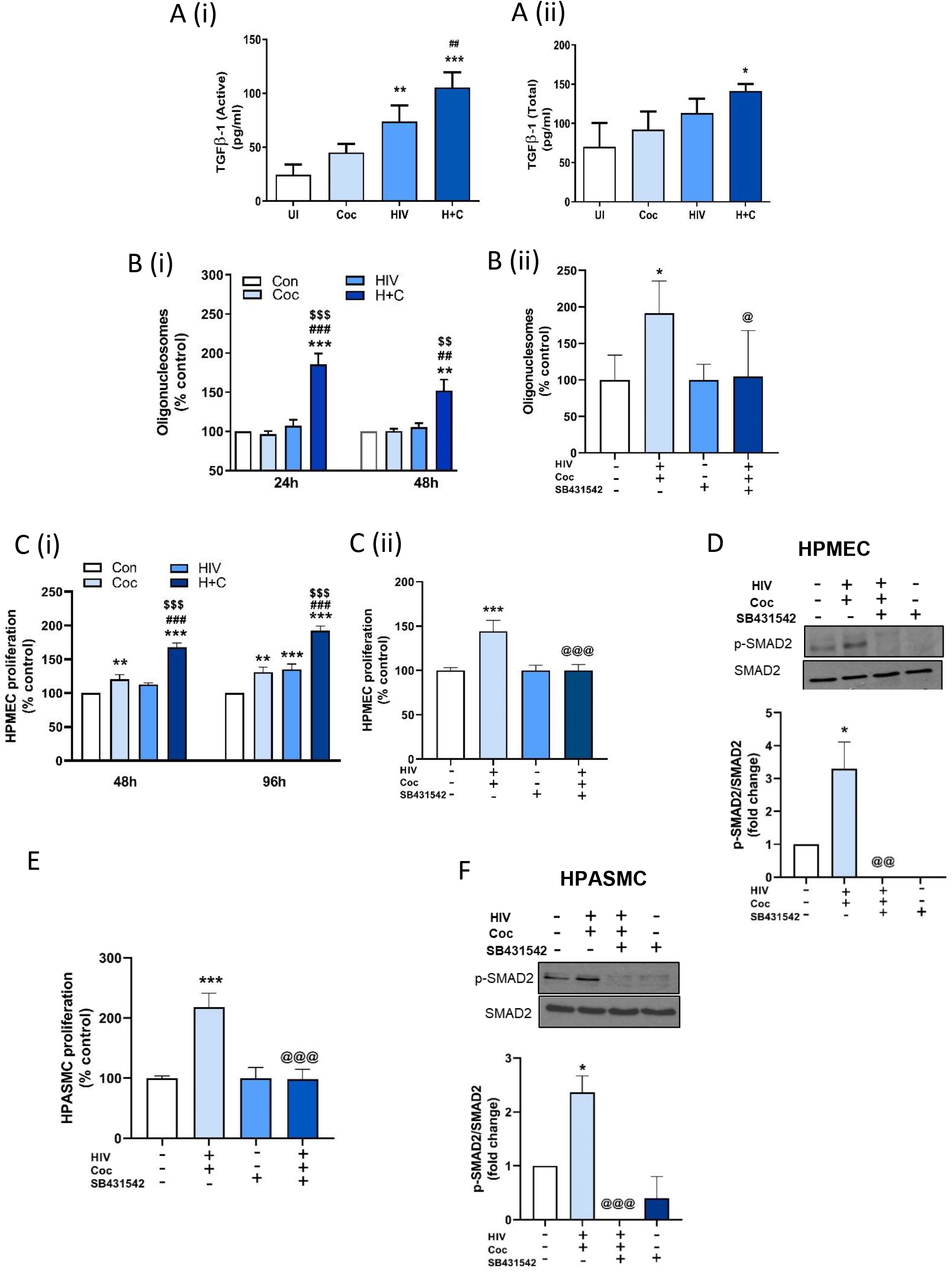
TGFβ-1 levels are up-regulated in EVs from HIV-infected monocyte-derived macrophages (MDMs) on treatment with cocaine resulting in enhanced MDM-EV- mediated endothelial dysfunction and smooth muscle hyperplasia. **A**, EVs isolated from supernatants of Monomac-1-derived macrophages infected with HIV-1_Bal_ for 4 days in the presence (H+C) and absence of cocaine (HIV) or un-infected MDMs treated with (Coc) or without cocaine (UI) were analyzed for active **(i)** and total **(ii)** TGFβ-1 levels by ELISA. **B**, HPMECs were treated with 4 µg MDM-EVs followed by cell death ELISA at 24 & 48h post EV addition in the absence (i) or at 48h post EV addition in the presence of TGFβ-R1 inhibitor (SB431542) (ii) **C**, MTS assay was performed on HPMECs at 48 or 96h post treatment with MDM-EVs in the absence (i) or at 48 h post EV treatment in the presence of SB431542 (ii)**. D**, HPMECs treated with H+C MDM-EVs in the presence and absence of SB431542 were lysed using RIPA buffer at 16 h post-treatment, followed by western blot analysis for phosphorylated and total SMAD2. The graphs represent the densitometry analysis of three independent experiments (mean±SEM). **E**, MTS assay was performed on HPASMCs treated with H+C MDM-EVs in the presence and absence of SB431542 at 48h post-treatment. **F**, Western blot analysis of HPASMCs for phosphorylated and total SMAD2 at 1h post treatment. For all inhibitor studies cell were pre-treated with SB431542 (10µM) for 30 min before the addition of MDM-EVs on cells. Values of experiments using MDM-EVs are average of 3 independent experiments. ***p< 0.001, **p< 0.01,*p< 0.05, vs control EVs; ### p< 0.001, ## p< 0.01 vs Coc EVs; $$$p< 0.001, $$ p< 0.01 vs HIV EVs and @@@ p< 0.001, @@p< 0.01, @p< 0.05 vs H+C EVs.

Exposure of HPMECs to H+C MDM-EVs lead to a significant increase in apoptosis at 24h and 48h post-treatment as compared to cells treated with EVs derived from un-infected (Con), HIV-infected (HIV) or cocaine-treated (Coc) MDMs (Fig. 4Bi and Supplementary fig. II- B). Besides, a significant endothelial proliferation of apoptosis resistant HPMECs was observed on treatment with H+C MDM-EVs when compared with all other groups at 48h and 96h (Fig. 4Ci & Supplementary figure II-C). However, addition of MDM-EVs in the presence of TGFβ-R1 inhibitor resulted in significant abrogation of H+C MDM-EV mediated induction of endothelial apoptosis (Fig. 4Bii) and proliferation (Fig. 4Cii) along with the prevention of SMAD2 activation (Fig. 4D).

Similarly, H+C MDM-EV mediated hyper-proliferation of smooth muscle cells was observed to be TGF-β dependent (Fig. 4E). In parallel, increased expression of phosphorylated-SMAD2 on treatment with H+C MDM-EVs attenuated on pretreatment of cells with TGFβ-R1 inhibitor (Fig. 4F).

### Right ventricle dysfunction in rats treated with HIV-cocaine MDM-EVs

To see the *in-vivo* effect of H+C MDM-EVs in the development of PH, we first confirmed the cellular uptake of PKH67-labeled human MDM-EVs by rat cells (Supplementary figure III-A) and *in vivo* bio- distribution of intravenously injected DiR-labeled human MDM-EVs in rat lungs (Supplementary figure III-B). Next, we investigated the effect of H+C MDM-EVs on hemodynamics in WT rats and whether they can further potentiate the RV dysfunction in our previously established non-infectious HIV-Tg rat PAH model (10). We observed a significant increase in the right ventricular systolic pressure (RVSP) (p = 0. 003; unpaired t-test) in WT rats given H+C-EVs compared to rats given uninfected MDM-EVs (Con-EVs). However, comparison between multiple groups (Figure 5B) revealed significant increase in RVSP in only HIV-Tg rats treated with either Con-EVs or H+C-EVs when compared to WT-Con EV group. Interestingly, RVSP in HIV-Tg rats given H+C-EVs exhibited higher trend compared to HIV-Tg rats given Con-EVs and a significant increase compared to WT rats administered H+C-EVs. But assessment of Fulton Index (RV/LV+Septum) showed significant RV hypertrophy only in HIV-Tg rats that were given Con-EVs and only increased trend in HIV-Tg rats given H+C-EVs when compared to the WT groups (Fig. 5C). The trichrome staining of right ventricles exhibited maximum fibrosis and collagen deposition in HIV-Tg rats administered H+C-EVs (Fig. 5E) and no significant differences were observed in the mean arterial pressure (MAP) among all four groups (Fig. 5D). Furthermore, HIV-Tg rats treatment with H+C-EVs resulted in significant increased levels of serum endothelin-1 (ET-1) and tumor necrosis factor-α (TNF-α) along with increased trend of C-reactive protein (CRP) compared to WT and HIV-Tg rats treated with Con-EVs (Fig. 5F). However, no differences were observed in IL-6 levels (Fig. 5F). Both ET-1 and TNF-α levels had a significant positive correlation with RVSP as shown in Figure 5G.

**Figure 5:**
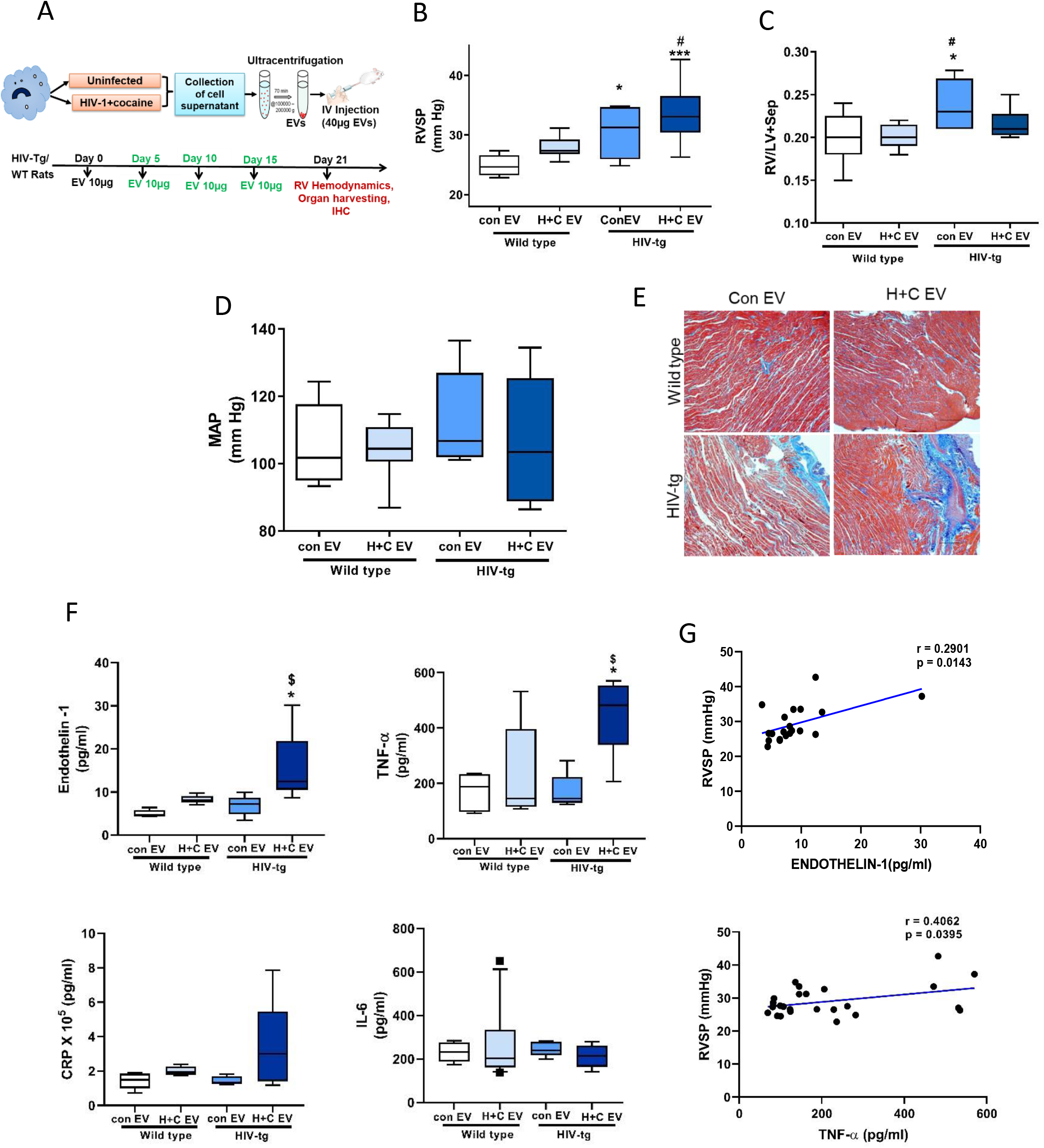
Hemodynamic measurements, right ventricle fibrosis and biomarkers of PH in rats injected with HIV-infected cocaine treated macrophage-derived EVs. **A**, Figure showing timeline for the administration of HIV-infected cocaine treated MDM-EVs (H+C EV) in WT/HIV-Tg rats. HIV-Tg/ WT rats injected with EVs from uninfected MDMs (Con EV) were used for comparison. Rats were intravenously injected 4 doses of 10µg EVs each at 5 day intervals. **B-D**, Box and whisker plots showing right ventricle systolic pressure (RVSP) (B) Fulton Index (C) and Mean arterial pressure (MAP) measurements (D). **E**, Representative micrographs show RV fibrosis after Masson’s trichrome staining on formaldehyde-fixed, paraffin embedded RV sections. Magnification 4X (Scale: 200µm). **F**, ELISAs were performed to assess serum levels of endothelin-1, TNF-β; C-reactive protein (CRP) and IL-6. **G**, Graphs showing correlation of RVSP with Endothelin-1 and TNF-β serum levels in rats. n = 6–8 rats per group. Box and whisker plots show the median value and 10–90 percentiles. ***p< 0.001, *p< 0.05 vs. WT-con EV; #p< 0.05 vs. WT-H+C EV, $p< 0.05 vs. HIV-Tg-con EV.

### Myocardial injury in rats treated with HIV-cocaine MDM-EVs

Given that we observed no further increase in Fulton index in HIV-Tg rats on administration of H+C-EVs compared to Con-EVs, we separately compared RV and LV+septum mass with body weight. Though the body weight remained constant in WT rats or HIV-Tg rats injected with Con-EVs or H+C-EVs (Supplementary figure IVA), both WT and HIV-Tg rats that were administered H+C-EVs had decreased RV and LV+septum mass (Fig. 6A-B). However, the effect on RV mass was far less compared to decrease in LV+septum mass (22–30% RV vs. 60–70% LV) among H+C-EV treated rats. This was associated with significantly increased caspase-3 expression in whole LV lysates (Fig. 6D), but not in RV lysates (Fig. 6C) from HIV-Tg rats treated with H+C-EVs when compared to rats treated with Con-EVs. Interestingly, significant increase in caspase-3 expression was observed in RV from WT rats given H+C-EVs compared to WT rats treated with Con-EVs (Fig. 6C). Decrease in both RV and LV cardiomyocyte size in HIV-Tg rats treated with H+C-EV compared to HIV-Tg rats given Con-EVs (Fig 6E-F) further suggested cardiac atrophy of HIV-Tg rats in response to treatment with H+C MDM-EVs. Finally, serum levels of cardiac Troponin-I (cTnI), an indicator of cardiomyocyte damage were increased in HIV-Tg animals given H+C-EVs compared to Con-EVs treated rats (Fig. 6G) and were found to positively correlate with RVSP (Fig. 6H).

**Figure 6:**
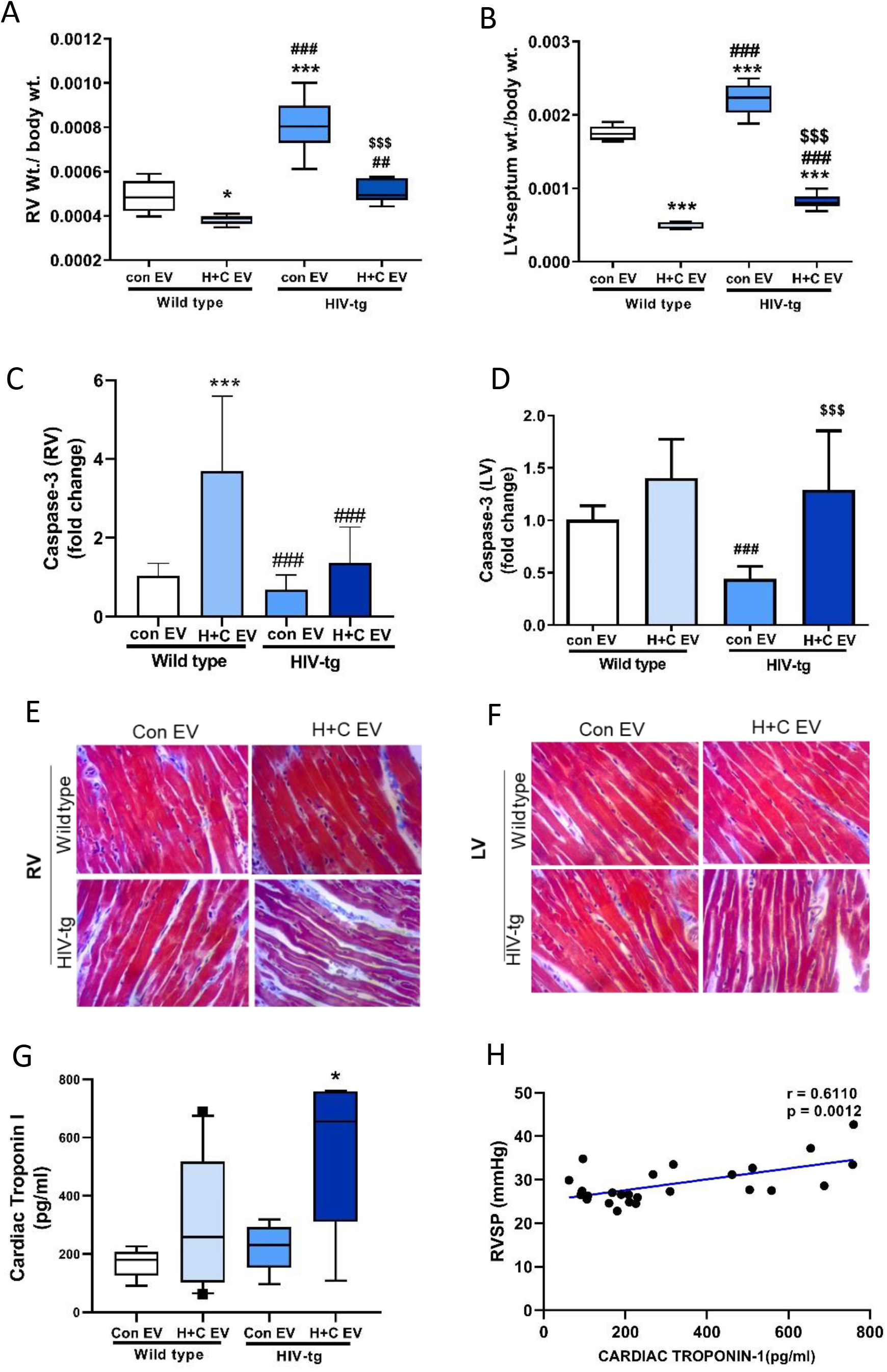
Analysis of cardiac atrophy in rats injected with EVs from HIV-infected and cocaine treated macrophages. **A-B**, Ratio of right ventricle (RV) weight to total body weight (A) and left ventricle (LV)+ septum weight /body weight. (B) for WT and HIV-Tg rats injected with EVs from uninfected monocyte derived macrophages (MDM) (Con-EV) or EVs from HIV-infected and cocaine treated MDMs (H+C-EV). Values are from n = 7–8 rats per group. **C-D**, Quantitative RT-PCR analysis of caspase-3 in the RV (C) and LV (D) tissues from rats (n = 6/group). The values are expressed as fold change. **E-F**, Representative micrographs showing RV (E) and LV (F) cardiomyocytes from all 4 groups after Masson’s Trichrome staining. Magnification 40X, Scale: 100µm. **G**, ELISA was performed to check the levels of cardiac troponin-1 in rat serum from all groups. Values are from n = 7–8 rats per group **H**, Graph showing correlation of RVSP with cardiac troponin-1 levels in rats. For box and whisker plots boxes represent median and IQR, whiskers show 10–90 percentile. ***p< 0.001, *p< 0.05 vs WT-Con EVs; ###p< 0.001, ##p< 0.01 vs WT-H+C EVs, $$$p< 0.001 vs HIV-Tg-Con EVs.

### Enhanced pulmonary endothelial injury and smooth muscle cell hyperplasia in rats administered with HIV-cocaine MDM-EVs

Staining of lung sections for vWF and α-SMA indicated enhanced smooth muscle positivity (red fluorescence) in the pulmonary vessels of HIV-Tg rats injected with Con-EVs or H+C-EVs as compared to both groups of WT rats. Increased SMA-positive pulmonary vessels was also found in WT rats injected with H+C-EVs when compared with WT rats given Con-EVs (Fig. 7A). Concordantly, RPASMCs from HIV-Tg rats injected with H+C-EVs exhibited significant hyper-proliferation as early as day 4 of serum-stressed conditions compared to HIV-Tg or WT rats given Con-EVs (Fig. 7B). At day 6 of serum-starvation, RPASMCs from both HIV-Tg rats and WT rats given H+C-EVs exhibited hyper-proliferation when compared to WT Con-EV treated rats. Simultaneously, RPASMCs from HIV-Tg rats showed less apoptotic cell death compared to WT rats at 48h of serum-starvation (Supplementary figure IV-B). These findings corresponded with increased TGFβ- R1activation in RPASMCs (Fig. 7C) and downstream p-SMAD2 expression (Supplementary figure IV-C) from H+C-EV treated rats compared to rats given Con-EVs.

**Figure 7:**
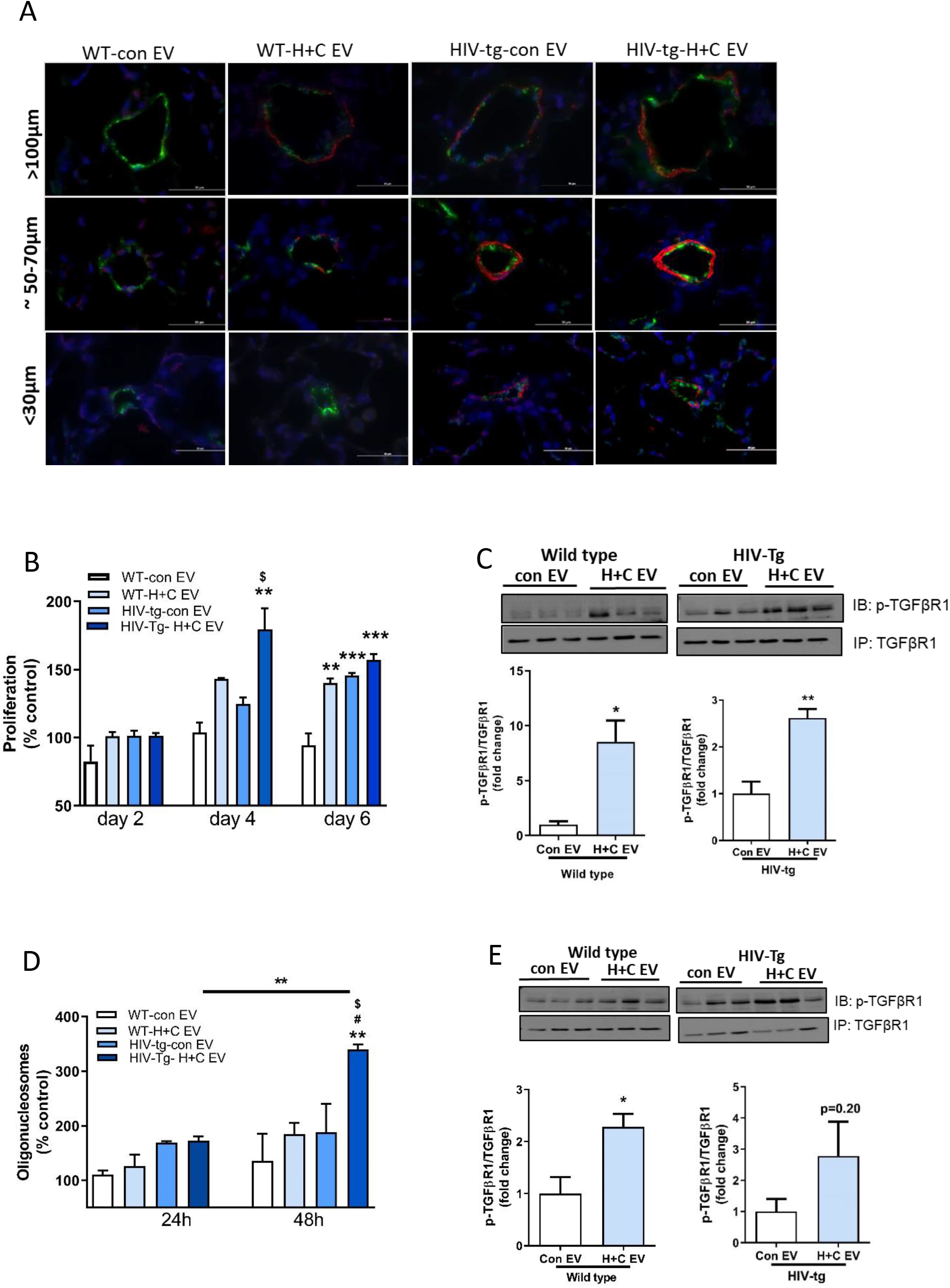
Pulmonary smooth muscle proliferation and endothelial injury in lungs from rats injected with HIV-infected cocaine treated MDM-EVs. **A**. Representative micrographs showing immunofluorescence staining of von Willebrand factor (vWF) (green) and β-smooth muscle actin (β-SMA)(red) in pulmonary vessels of size > 100µm, 50–70µm and < 30µm in all 4 groups (Scale bar = 50µm). **B**, Graph showing percent proliferation of rat pulmonary arterial smooth muscle cells (RPASMCs) isolated from all 4 groups. RPASMCs were cultured in 96-well plates and MTS assay was performed at day 2, 4 and 6 to assess the cell proliferation. **C**, Total extract of RPASMCs (n = 3/group) was immune-precipitated (IP) using TGFβ-R1 antibody followed by western blot (IB) and detection of phosphorylated and total levels of TGFβ-R1. Graphs represent densitometric analysis of n = 3 animals per group. **D**, Cell death ELISA on rat pulmonary microvascular endothelial cells (RPMECs) isolated from all 4 groups. RPMECs isolated from left lung lobe were cultured in 96-well plates and the oligonucleosome ELISA was performed at 24 and 48h to assess the cell apoptosis. **E**, IP-western blot and densitometric analysis showing phosphorylated and total levels of TGFβ-R1 in RPMECs. Values are Mean±SEM of n = 3/group ***p< 0.001, **p< 0.01, *p< 0.05 vs WT-con EVs, #p< 0.05 vs WT-H+C EVs, $p< 0.05 vs HIV-Tg-Con EVs.

Contrary to RPASMCs, RPMECs from HIV-Tg rats given H+C-EVs demonstrated significantly higher apoptosis in response to serum-starvation when compared with other groups (Fig. 7D) and exhibited high activated caspase-3 (Supplementary figure IV-D). In addition, these ECs exhibited less survivability to serum-starvation (Supplementary figure IV-E) and increased TGFβ-R1 (Fig. 7E) and SMAD2 activation (Supplementary figure IV-F).

### Pharmacological inhibition of TGFβ-R1 resulted in attenuation of HIV-cocaine MDM-EVs mediated cardiopulmonary dysfunction

For *in-vivo* inhibition of TGF-β downstream signaling, we treated rats with GW788388 that is considered to be more potent TGFβ-R1 (Alk-5) inhibitor compared to SB431542 without having any non-specific effects on bone morphogenetic protein receptor (BMPR)-2 signaling. Animals from both WT and HIV-Tg groups pre-treated with GW788388 showed improvement in H+C-EV mediated increase in RVSP (Fig. 8A). Echocardiography performed on this set of rats revealed an increase in RV end-diastolic area (EDA) after 21days of treatment with H+C-EVs whereas blocking of TGF-βRI prevented this increase (Fig. 8B) as also shown by a significant drop in differences between RV/LV EDA assessed at day 21 and at day 0 (Fig. 8C). Correspondingly, significant increase in the difference between RV/LV ejection fraction (EF) pre- and post- treatment was observed in WT and HIV-Tg rats treated with H+C-EVs in the presence of inhibitor compared to H+C-EVs rats treated with vehicle (Fig. 8D). A parallel mitigation of H+C-EV mediated increase in the levels of ET-1, TNF-α and cardiotroponin I was also observed on treatment with GW7883888 (Fig. 8E).

**Figure 8:**
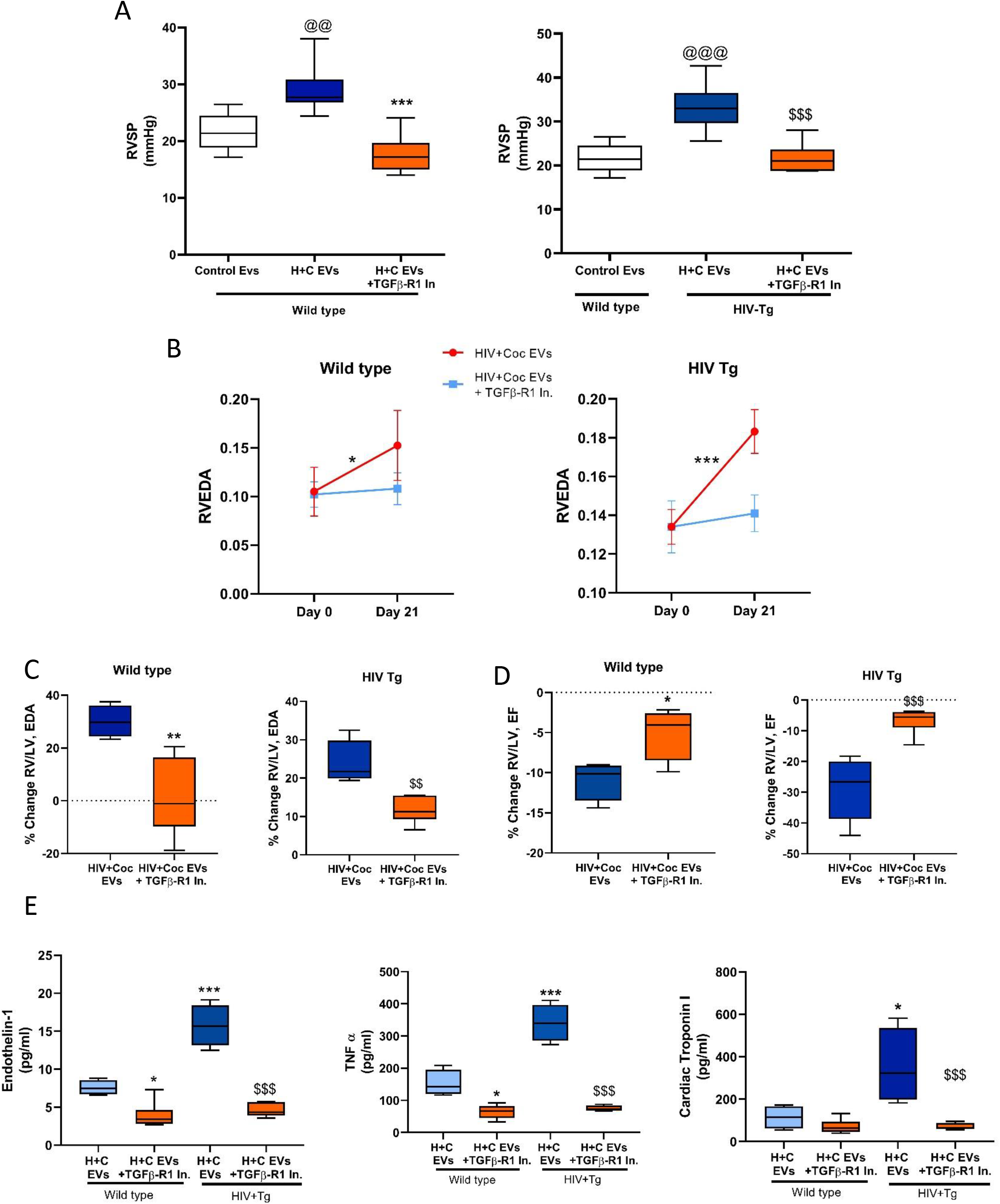
Attenuation of HIV-infected cocaine treated MDM-EVs mediated cardio-pulmonary dysfunction in the presence of TGFβ-RI inhibitor. HIV-Tg or wild type (WT) rats were intravenously injected 4 doses of H+C MDM-EVs, 10µg each at 5 days interval in the presence of treatment with TGFβ-Receptor inhibitor: GW788388 (1mg/kg/day) or vehicle, once orally for 21 days (n = 5/group). **A**, Right ventricle systolic pressure (RVSP) in WT and HIV-Tg rats given H+C EVs with and without inhibitor in comparison to WT rats treated with control EVs. **B**. Right ventricle end diastolic area (RVEDA) at day 0 and day 21 for WT and HIV-Tg rats given H+C-EVs in the presence or absence of GW788388. **C-D**. Percentage change in RV/LV, EDA (C) and Ejection Fraction (EF) in rats on treatment with or without GW788388 (D) **E**. ELISAs were performed to assess serum levels of endothelin-1, TNF-β and Cardiac troponin-I. Box and whisker plots show the median value and 10–90 percentiles. @p< 0.05, @@@p< 0.001 vs control EVs wild type, *p< 0.05, **p< 0.01, ***p< 0.001 vs wild type H+C EVs, $$p< 0.01, $$$p< 0.001 vs HIV-Tg H+C EVs

## DISCUSSION

In this study, we report a higher number of TGF-β loaded EVs in the plasma from HIV-PH individuals compared to plasma from HIV-non PH individuals, suggesting its potential as a biomarker for HIV-PH. We showed that the EVs from plasma of HIV-infected cocaine users and from HIV-infected MDMs treated with cocaine augment pulmonary endothelial injury and smooth muscle hyperplasia via activation of TGF-β1 signaling. We show for the first time that the EVs derived from HIV-infected cocaine MDMs can cause pulmonary vascular remodeling and promote the development of PH *in-vivo*.

PAH has been well-established as a secondary complication to HIV infection. The link between HIV infection leading to pulmonary vascular remodeling (6, 9) and further contribution of IVDU in exacerbating the development of PAH has been well-established (26-28). Viral proteins show an additive effect with illicit drugs such as cocaine and opioids in potentiating pulmonary vascular remodeling and development of PH in infectious non-human primate or non-infectious HIV-transgenic rat models (9, 10, 29). Recently, EVs have been recognized as novel communicators between cells and thereby contributing to disease pathogenesis. Importantly, we and others have shown that the level and content of EVs released by immune cells changes with virus replication (15, 19). Many studies have reported increased levels of plasma- or platelet-derived EVs in patients with idiopathic, heritable or connective tissue disease-associated PAH (15, 21, 30). Here, we show increased number of small-sized PEVs in HIV-PH individuals not only compared to un-infected individuals, but also significantly higher compared to HIV-infected individuals without PH. Furthermore, consistent with our earlier findings of augmented pulmonary vascular damage in response to the dual hit of drug abuse and HIV-infection, we observed higher number of PEVs in HIV-infected cocaine users compared to HIV-infected non-drug users or un-infected cocaine users.

Inflammation is the hallmark feature of all forms of PAH with increased infiltration of inflammatory cells such as macrophages and T-cells surrounding the pulmonary vessels (7, 8, 12). We previously demonstrated significant perivascular inflammation in morphine-treated simian human immunodeficiency virus (SHIV) infected macaques as well as in HIV-Tg rats exposed to cocaine (9). Similarly increased numbers of pro-inflammatory and pro-fibrotic monocytes have been observed in the peripheral blood (31) of SHIVnef- or SIV-infected macaques. Although these studies suggest the association of inflammation with HIV-PAH, studies defining the direct role of inflammatory cells in the development of the disease are limited. Our current study demonstrates the direct effect of EVs derived from HIV-infected monocyte derived macrophages in causing cardio-pulmonary dysfunction.

Macrophage depletion has been reported to prevent the development of PH in animals (32). Recent study by Florentin et al reported that increased number of blood monocytes during PAH get recruited to lung perivascular spaces and differentiate into inflammatory macrophages responsible for pulmonary vascular remodeling (33). Our pivotal findings suggest that the altered cargo within the circulating EVs derived from H+C-MDMs are able to cause EC injury and SMC proliferation, consequently leading to the development of PH in WT rats. In addition, these H+C MDM-EVs potentiated the vascular damage in the HIV Tg-cocaine rat model of PAH (10) along with increasing serum levels of TNF-α and endothelin-1 known to affect the vascular cell function and facilitate the development of PH (34) (35). Previous studies on PEVs from monocrotaline-induced PAH mice have shown to cause PAH when injected into healthy mice (15, 16). However, EVs are known to transfer their contents not only to different cell types, but also across different species (36, 37). Our data show that injecting HIV-infected human cell-derived EVs into HIV-Tg rats can be used as a tool to delineate the relationship between immune activation/inflammation and non-AIDS comorbidities in this non-infectious HIV-Tg rat model.

Although administration of H+C-EVs in WT and HIV-Tg rats resulted in increased RVSP, analysis of right and left ventricles indicated a significant decrease in LV+septum mass. LV dysfunction with decreased left ventricular mass index has been reported earlier in HIV-infected individuals (38). Changes in rat LV mass can be explained due to inter-ventricular dependence as a result of RV overload associated with high pulmonary vascular resistance (PVR) (39), (40, 41). Other possibility could be the direct effect of H+C EVs on cardiomyocytes leading to increased apoptosis and atrophy. We observed increased levels of circulating cardiac troponin I, CRP and TNF-α indicative of cardiac injury and inflammation in response to H+C- EV treatment, but without an increase in pro-inflammatory IL-6.

TGF-β1, a growth factor and pro-inflammatory cytokine is known to play an important role in pulmonary vascular remodeling and PH (22, 42, 43). We found that cocaine exposure not only augmented the levels of TGF-β in the PEVs from PLWH and EVs from HIV-infected MDMs, but also increased the levels in EVs independent of infection. Pre-treatment of cells with TGF-β receptor inhibitor resulted in attenuation of H+C PEV/MDM-EV mediated pulmonary vascular dysfunction. Recent reports demonstrated that TGFβ-1-associated exosomes promote more potent and sustained activation of downstream signaling compared to free TGF-β1 (25). Our findings showing TGF-β dependent escalation of cardio-pulmonary dysfunction in H+C-EV treated rats further emphasizes its significance in EV-mediated pathological changes. However, TGF-β1 may not be the only molecule and additional cargo molecules including other proteins and miRNAs in these EVs may also contribute to pulmonary vascular remodeling *in-vivo*. We earlier reported that increased levels of miR-130 in H+C-MDM EVs mediates pro-survival signaling in HPASMCs (21). However, we were unable to detect miR-130 in PEVs from any of the groups tested. Nevertheless, higher TGF-β levels in the plasma-EVs from HIV-PH individuals compared to HIV-infected non-PH individuals and its significant correlation with PASP and ability to cause EC/SMC proliferation highlight the importance of TGF-β-rich EVs in PH development.

The exoEasy method that we used to isolate PEVs has recently been reported to be most practical, faster and reproducible over traditional approaches to isolate good quality EVs from small volume serum or plasma samples for the downstream application (44). Nevertheless, our study includes limitations of analyzing only total number of plasma EVs without defining the nature and origin of PEVs. Other limitations include a small sample size and lack of confirmation of PH using right heart catheterization in PLWH. Further investigations using a larger cohort are needed to better characterize the EVs and confirm the correlation of TGFβ-loaded EVs with the clinical parameters of PH to use it as a circulatory biomarker of HIV-PH.

In summary, we report for the first time the direct effect of HIV-infected macrophage derived small EVs on pulmonary vascular modeling and PH development via modulation of TGF-β signaling in pulmonary SMCs and ECs. Most importantly, this is the first study that correlates increased number of TGF-β1 linked PEVs with the presence of PH in HIV-infected individuals. It will be interesting to see if these TGF-β rich plasma-EVs are also high in other forms of PH. Further investigations of these EVs to elucidate their potential as biomarker and therapeutic target for PH are warranted.

## Data Availability

The datasets generated during the current study will be available from the corresponding author on reasonable request.

## ACKNOWLEDGMENTS

Authors are grateful to Dr. Ghazwan Butrous, University of Kent, Canterbury, UK for providing invaluable feedback and critical evaluation of this manuscript. We acknowledge Julie Allen, Department of Molecular and Integrative Physiology, KUMC for helping in animal surgery; Michael Wulser, Department of Internal Medicine, KUMC for morphometric analysis and; Pranjali Dalvi and Himanshu Sharma, Department of Internal Medicine, KUMC for initial standardization of *in-vitro* EV uptake and proliferation/apoptosis experiments. We also acknowledge Electron Microscope Research Laboratory (NIH/NIGMS COBRE grant P20GM104936 and NIH grant 1S10RR027564) for TEM analysis.

## REFERENCES

1. Triplette M, Crothers K, Attia EF. Non-infectious Pulmonary Diseases and HIV. Curr HIV/AIDS Rep 2016; 13: 140–148.

2. Quezada M, Martin-Carbonero L, Soriano V, Vispo E, Valencia E, Moreno V, de Isla LP, Lennie V, Almeria C, Zamorano JL. Prevalence and risk factors associated with pulmonary hypertension in HIV-infected patients on regular follow-up. AIDS 2012; 26: 1387–1392.

3. Harter ZJ, Agarwal S, Dalvi P, Voelkel NF, Dhillon NK. Drug abuse and HIV-related pulmonary hypertension: double hit injury. AIDS 2018; 32: 2651–2667.

4. Mermis J, Gu H, Xue B, Li F, Tawfik O, Buch S, Bartolome S, O'Brien-Ladner A, Dhillon NK. Hypoxia-inducible factor-1 alpha/platelet derived growth factor axis in HIV-associated pulmonary vascular remodeling. Respir Res 2011; 12: 103.

5. Almodovar S. The complexity of HIV persistence and pathogenesis in the lung under antiretroviral therapy: challenges beyond AIDS. Viral Immunol 2014; 27: 186–199.

6. Fitzpatrick M, Crothers K, Morris A. Future directions: lung aging, inflammation, and human immunodeficiency virus. Clin Chest Med 2013; 34: 325–331.

7. Savai R, Pullamsetti SS, Kolbe J, Bieniek E, Voswinckel R, Fink L, Scheed A, Ritter C, Dahal BK, Vater A, Klussmann S, Ghofrani HA, Weissmann N, Klepetko W, Banat GA, Seeger W, Grimminger F, Schermuly RT. Immune and inflammatory cell involvement in the pathology of idiopathic pulmonary arterial hypertension. Am J Respir Crit Care Med 2012; 186: 897–908.

8. Rabinovitch M, Guignabert C, Humbert M, Nicolls MR. Inflammation and immunity in the pathogenesis of pulmonary arterial hypertension. Circ Res 2014; 115: 165–175.

9. Spikes L, Dalvi P, Tawfik O, Gu H, Voelkel NF, Cheney P, O'Brien-Ladner A, Dhillon NK. Enhanced pulmonary arteriopathy in simian immunodeficiency virus-infected macaques exposed to morphine. Am J Respir Crit Care Med 2012; 185: 1235–1243.

10. Dalvi P, Spikes L, Allen J, Gupta VG, Sharma H, Gillcrist M, Montes de Oca J, O'Brien-Ladner A, Dhillon NK. Effect of Cocaine on Pulmonary Vascular Remodeling and Hemodynamics in Human Immunodeficiency Virus-Transgenic Rats. Am J Respir Cell Mol Biol 2016; 55: 201–212.

11. Soon E, Holmes AM, Treacy CM, Doughty NJ, Southgate L, Machado RD, Trembath RC, Jennings S, Barker L, Nicklin P, Walker C, Budd DC, Pepke-Zaba J, Morrell NW. Elevated levels of inflammatory cytokines predict survival in idiopathic and familial pulmonary arterial hypertension. Circulation 2010; 122: 920–927.

12. Tuder RM, Groves B, Badesch DB, Voelkel NF. Exuberant endothelial cell growth and elements of inflammation are present in plexiform lesions of pulmonary hypertension. Am J Pathol 1994; 144: 275–285.

13. Osada-Oka M, Shiota M, Izumi Y, Nishiyama M, Tanaka M, Yamaguchi T, Sakurai E, Miura K, Iwao H. Macrophage-derived exosomes induce inflammatory factors in endothelial cells under hypertensive conditions. Hypertens Res 2017; 40: 353–360.

14. Tang N, Sun B, Gupta A, Rempel H, Pulliam L. Monocyte exosomes induce adhesion molecules and cytokines via activation of NF-kappaB in endothelial cells. FASEB J 2016; 30: 3097–3106.

15. Aliotta JM, Pereira M, Wen S, Dooner MS, Del Tatto M, Papa E, Goldberg LR, Baird GL, Ventetuolo CE, Quesenberry PJ, Klinger JR. Exosomes induce and reverse monocrotaline-induced pulmonary hypertension in mice. Cardiovasc Res 2016; 110: 319–330.

16. Aliotta JM, Pereira M, Amaral A, Sorokina A, Igbinoba Z, Hasslinger A, El-Bizri R, Rounds SI, Quesenberry PJ, Klinger JR. Induction of pulmonary hypertensive changes by extracellular vesicles from monocrotaline-treated mice. Cardiovasc Res 2013; 100: 354–362.

17. Amabile N, Heiss C, Real WM, Minasi P, McGlothlin D, Rame EJ, Grossman W, De Marco T, Yeghiazarians Y. Circulating endothelial microparticle levels predict hemodynamic severity of pulmonary hypertension. Am J Respir Crit Care Med 2008; 177: 1268–1275.

18. Bakouboula B, Morel O, Faure A, Zobairi F, Jesel L, Trinh A, Zupan M, Canuet M, Grunebaum L, Brunette A, Desprez D, Chabot F, Weitzenblum E, Freyssinet JM, Chaouat A, Toti F. Procoagulant membrane microparticles correlate with the severity of pulmonary arterial hypertension. Am J Respir Crit Care Med 2008; 177: 536–543.

19. Chelvanambi S, Bogatcheva NV, Bednorz M, Agarwal S, Maier B, Alves NJ, Li W, Syed F, Saber MM, Dahl N, Lu H, Day RB, Smith P, Jolicoeur P, Yu Q, Dhillon NK, Weissmann N, Twigg Iii HL, Clauss M. HIV-Nef Protein Persists in the Lungs of Aviremic Patients with HIV and Induces Endothelial Cell Death. Am J Respir Cell Mol Biol 2019; 60: 357–366.

20. Chelvanambi S, Gupta SK, Chen X, Ellis BW, Maier BF, Colbert TM, Kuriakose J, Zorlutuna P, Jolicoeur P, Obukhov AG, Clauss M. HIV-Nef Protein Transfer to Endothelial Cells Requires Rac1 Activation and Leads to Endothelial Dysfunction Implications for Statin Treatment in HIV Patients. Circ Res 2019; 125: 805–820.

21. Sharma H, Chinnappan M, Agarwal S, Dalvi P, Gunewardena S, O'Brien-Ladner A, Dhillon NK. Macrophage-derived extracellular vesicles mediate smooth muscle hyperplasia: role of altered miRNA cargo in response to HIV infection and substance abuse. FASEB J 2018; 32: 5174–5185.

22. Kumar R, Mickael C, Kassa B, Gebreab L, Robinson JC, Koyanagi DE, Sanders L, Barthel L, Meadows C, Fox D, Irwin D, Li M, McKeon BA, Riddle S, Dale Brown R, Morgan LE, Evans CM, Hernandez-Saavedra D, Bandeira A, Maloney JP, Bull TM, Janssen WJ, Stenmark KR, Tuder RM, Graham BB. TGF-beta activation by bone marrow-derived thrombospondin-1 causes Schistosoma- and hypoxia-induced pulmonary hypertension. Nat Commun 2017; 8: 15494.

23. Dalvi P, Sharma H, Konstantinova T, Sanderson M, Brien-Ladner AO, Dhillon NK. Hyperactive TGF-beta Signaling in Smooth Muscle Cells Exposed to HIV-protein(s) and Cocaine: Role in Pulmonary Vasculopathy. Sci Rep 2017; 7: 10433.

24. Hemnes AR, Humbert M. Pathobiology of pulmonary arterial hypertension: understanding the roads less travelled. Eur Respir Rev 2017; 26.

25. Shelke GV, Yin Y, Jang SC, Lasser C, Wennmalm S, Hoffmann HJ, Li L, Gho YS, Nilsson JA, Lotvall J. Endosomal signalling via exosome surface TGFbeta-1. J Extracell Vesicles 2019; 8: 1650458.

26. Porter KM, Walp ER, Elms SC, Raynor R, Mitchell PO, Guidot DM, Sutliff RL. Human immunodeficiency virus-1 transgene expression increases pulmonary vascular resistance and exacerbates hypoxia-induced pulmonary hypertension development. Pulm Circ 2013; 3: 58–67.

27. George MP, Champion HC, Gladwin MT, Norris KA, Morris A. Injection drug use as a “second hit” in the pathogenesis of HIV-associated pulmonary hypertension. Am J Respir Crit Care Med 2012; 185: 1144–1146.

28. Janda S, Quon BS, Swiston J. HIV and pulmonary arterial hypertension: a systematic review. HIV Med 2010; 11: 620–634.

29. Dhillon NK, Li F, Xue B, Tawfik O, Morgello S, Buch S, Ladner AO. Effect of cocaine on human immunodeficiency virus-mediated pulmonary endothelial and smooth muscle dysfunction. Am J Respir Cell Mol Biol 2011; 45: 40–52.

30. Zhao L, Luo H, Li X, Li T, He J, Qi Q, Liu Y, Yu Z. Exosomes Derived from Human Pulmonary Artery Endothelial Cells Shift the Balance between Proliferation and Apoptosis of Smooth Muscle Cells. Cardiology 2017; 137: 43–53.

31. Schweitzer F, Tarantelli R, Rayens E, Kling HM, Mattila JT, Norris KA. Monocyte and Alveolar Macrophage Skewing Is Associated with the Development of Pulmonary Arterial Hypertension in a Primate Model of HIV Infection. AIDS Res Hum Retroviruses 2019; 35: 63–74.

32. Tian W, Jiang X, Tamosiuniene R, Sung YK, Qian J, Dhillon G, Gera L, Farkas L, Rabinovitch M, Zamanian RT, Inayathullah M, Fridlib M, Rajadas J, Peters-Golden M, Voelkel NF, Nicolls MR. Blocking macrophage leukotriene b4 prevents endothelial injury and reverses pulmonary hypertension. Sci Transl Med 2013; 5: 200ra117.

33. Florentin J, Coppin E, Vasamsetti SB, Zhao J, Tai YY, Tang Y, Zhang Y, Watson A, Sembrat J, Rojas M, Vargas SO, Chan SY, Dutta P. Inflammatory Macrophage Expansion in Pulmonary Hypertension Depends upon Mobilization of Blood-Borne Monocytes. J Immunol 2018; 200: 3612–3625.

34. Courboulin A, Tremblay VL, Barrier M, Meloche J, Jacob MH, Chapolard M, Bisserier M, Paulin R, Lambert C, Provencher S, Bonnet S. Kruppel-like factor 5 contributes to pulmonary artery smooth muscle proliferation and resistance to apoptosis in human pulmonary arterial hypertension. Respir Res 2011; 12: 128.

35. Selimovic N, Bergh CH, Andersson B, Sakiniene E, Carlsten H, Rundqvist B. Growth factors and interleukin-6 across the lung circulation in pulmonary hypertension. Eur Respir J 2009; 34: 662–668.

36. Dorronsoro A, Robbins PD. Regenerating the injured kidney with human umbilical cord mesenchymal stem cell-derived exosomes. Stem cell research & therapy 2013; 4: 39.

37. Zhou Y, Xu H, Xu W, Wang B, Wu H, Tao Y, Zhang B, Wang M, Mao F, Yan Y, Gao S, Gu H, Zhu W, Qian H. Exosomes released by human umbilical cord mesenchymal stem cells protect against cisplatin-induced renal oxidative stress and apoptosis in vivo and in vitro. Stem cell research & therapy 2013; 4: 34.

38. Martinez-Garcia T, Sobrino JM, Pujol E, Galvez J, Benitez E, Giron-Gonzalez JA. Ventricular mass and diastolic function in patients infected by the human immunodeficiency virus. Heart 2000; 84: 620–624.

39. Homsi R, Luetkens JA, Skowasch D, Pizarro C, Sprinkart AM, Gieseke J, Meyer Zur Heide Gen Meyer-Arend J, Schild HH, Naehle CP. Left Ventricular Myocardial Fibrosis, Atrophy, and Impaired Contractility in Patients With Pulmonary Arterial Hypertension and a Preserved Left Ventricular Function: A Cardiac Magnetic Resonance Study. J Thorac Imaging 2017; 32: 36–42.

40. Manders E, Rain S, Bogaard HJ, Handoko ML, Stienen GJ, Vonk-Noordegraaf A, Ottenheijm CA, de Man FS. The striated muscles in pulmonary arterial hypertension: adaptations beyond the right ventricle. Eur Respir J 2015; 46: 832–842.

41. Vizza CD, Lynch JP, Ochoa LL, Richardson G, Trulock EP. Right and left ventricular dysfunction in patients with severe pulmonary disease. Chest 1998; 113: 576–583.

42. Ismail S, Sturrock A, Wu P, Cahill B, Norman K, Huecksteadt T, Sanders K, Kennedy T, Hoidal J. NOX4 mediates hypoxia-induced proliferation of human pulmonary artery smooth muscle cells: the role of autocrine production of transforming growth factor-{beta}1 and insulin-like growth factor binding protein-3. Am J Physiol Lung Cell Mol Physiol 2009; 296: L489–499.

43. Yang S, Banerjee S, Freitas A, Cui H, Xie N, Abraham E, Liu G. miR-21 regulates chronic hypoxia-induced pulmonary vascular remodeling. Am J Physiol Lung Cell Mol Physiol 2012; 302: L521–529.

44. Stranska R, Gysbrechts L, Wouters J, Vermeersch P, Bloch K, Dierickx D, Andrei G, Snoeck R. Comparison of membrane affinity-based method with size-exclusion chromatography for isolation of exosome-like vesicles from human plasma. J Transl Med 2018; 16: 1.

